# Efficacy of Low Level Laser Therapy on The Post Midline laparotomy Wound Pain in upper and lower Gastrointestinal (GI) Surgical Procedure : a randomized clinical trial

**DOI:** 10.1101/2024.04.24.24306282

**Authors:** Madadi Mohammad, Heydari Soleiman, Hashemi Seyed Ebrahim, Raei Mehdi, Gholizadeh Hamed

## Abstract

**Introduction:** Painkiller drugs play an important role in reducing pain after surgery. These drugs might have unavoidable side effects and by the identification of new side effects, the needs for non-drug agents have been increased gradually. Therefore we decided to investigate the effect of low-power laser therapy on pain control after surgery.

**Materials and Methods:** In this study, 106 patients as candidate for elective gastrointestinal tract surgery after exclusion 6 case were divided into two groups with 50 patients after being randomized. The intervention group and control group underwent low level laser therapy and placebo by daily manner respectively after surgery. Then both groups were evaluated and compared in terms of pain intensity and amount of pethidine consumption.

**Results:** There was no significant difference between two groups in terms of average age, sexual frequency and perioperative inflammatory factors and skin complications. The average pain intensity at 0, 24, 48, 72, 96 hours after surgery was 7.2, 5.3, 4.2, 3.66, 2.44 for the intervention group and it was 7.14, 6.3, 5.3, 4.3, 2.7 for the control group respectively. Also, the average use of pethidine was evaluated in 24-hour intervals until the fourth day, which was 53, 29, 10, 5.5 mg for the intervention group and 54, 37.5, 22.5, 8.5 mg for the control group respectively. At the first 24 hours and 96 hours after the operation, difference of pain intensity and pethidine consumption between the two groups was not significant, but in period 24-96 hours after surgery, the pain intensity and also the average pethidine consumption significantly decreased in the intervention group compared to the control group.

**Conclusion:** The findings of this study showed that the low level laser therapy can be used to control the pain and can be proposed as an alternative method to painkillers drugs.

## Introduction

With the increase in the number of surgeries performed in various fields, the need to reduce hospitalization time and return patients to daily activities as soon as possible is one of the basic concerns at the level of treatment planning and hospital management (1). Acute pain after surgery consists of two types of pain processes: somatic pain and visceral pain; Somatic pain plays a more important role, and visceral pain is less important in causing acute pain after surgery due to the fact that it is mainly the result of stretching, tearing or inflammation (2).The main cause of somatic pain is damage to the surface tissues and sensory nerves during surgical incision, and of course, inflammation can intensify this process (2). Acute pain after surgery interferes with the physiological systems of various organs of the body and causes their dysfunction and causes mortality and morbidity and increases hospitalization time. For this reason, using methods that are effective in reducing the hospitalization time and reducing the pain of patients and their early recovery; It will be very helpful (3). One of these methods, which is referred to as a non-invasive treatment without pain or complications; Cool Laser or Low Level Laser light is used. In this method, Non-Thermal light photons are used to the desired location (3). In general, the use of lasers in medicine is divided into the following categories according to laser power (4):

1. Neutral reactions: during these reactions, biological processes do not change in reaction with light, and this effect is used in the manufacture of diagnostic devices.
2. Destructive reactions: In these reactions, the photo physical effects of light on living tissue lead to heat generation and tissue destruction, and these reactions are used in surgery.
3. Photochemical reactions: the energy absorbed by the living tissue leads to the activation of a series of biochemical processes in the cell and finally the process of biological construction takes place in the cell. These effects are caused by non-thermal photons in the tissue.

The analgesic effects of low-power laser therapy are due to the following mechanisms (4):

1. Increasing pain threshold.
2. Reducing the secretion of chemicals such as histamine and acetylcholine.
3. Reduction of bradykinin synthesis.
4. Increase in micro vascular flow followed by reduction of edema.
5. Increasing the activity of acetyl cholinesterase and preventing the accumulation of acetylcholine.
6. Reducing the production of substance P, that causes faster transmission of pain impulses.

Due to the fact that after abdominal surgery, chest and abdominal movements cause a lot of pain in patients, and this itself leads to a decrease in patient mobility, as well as a decrease in the depth of breathing and lack of coughing, and several complications such as DVT, PTE, pulmonary atelectasis, decrease in arterial oxygenation and ileus and finally increase the length of hospitalization of the patient. It also causes severe stimulation of the sympathetic nervous system, which can have irreparable side effects in patients with heart and lung problems. To control the pain, different drug methods are used. each of these drugs is associated with many side effects. In different studies, different laser therapy protocols with different number of sessions have been used. Therefore, we have planned our study with the low effective dose and the low number of sessions; to demonstrate the effectiveness of low level laser therapy and facilitate its use as a routine treatment protocol.

## Materials and Methods

Considering the first type error equal to 5% and the second type error equal to 20%, the power of the study is equal to 80%, the standard deviation is equal to 5, and the minimum average pain score difference between the two groups is equal to 3 (the average effect size is equal to 0.6); the number of samples for each group according to www.openepi.com should be considered at least 44 people, taking into account the possibility of dropping samples during the study, primary sample size include 106 people at first. 106 Patients with age range of 30 to 60 years and have undergone midline laparotomy for the purpose of elective upper and lower gastrointestinal surgery from 22 October 2022 until 19 march 2023 in Tehran’s Baqiyatallah hospital entered to study except following cases:

1. Patients who did not consent to participate in the study at any stage.
2. Patients whose surgical wounds are infected for any reason.
3. Patients who had another serious medical problem (ASA III-VI) or had psychotic disorders or drug addiction.

Finally after exclusion some ones and obtaining written inform consent as it been described in next part; 100 Patients were divided into control and intervention groups of 50 people by www.randomizer.org in double blinded and simple randomly manner. In both groups, preliminary laboratory tests were performed before and on the first and third day after the operation in terms of WBC, ESR and CRP. In the process of anesthetizing the patients, initial pretreatment with fentanyl 2 μg/kg and midazolam 0.03 mg/kg, induction of general anesthesia with propofol 2mg/kg, lidocaine 1mg/kg and atracurium 0.5mg/kg were performed. After about 3 minutes of ventilation with a suitable face mask and after complete muscle relaxation, airway intubation was performed with tracheal tube number 8 fr for men and 7.5 fr for women and after fixing the tracheal tube in the right place, anesthesia was maintained Intravenous and inhaled drugs (N_2_O 50% + _2_O 50% + isoflurane 1-1.5%) continued. At the end of the surgery, the wound was dressed and after proper breathing, the tracheal tube was removed and the patients were transferred to recovery. In addition to the usual painkillers (30 mg ketorolac as an intravenous infusion every 12 hours); the intervention group were received low level laser therapy from about 12-18 hours after the operation until the third day after the operation with 24-hour intervals (3 times); And the control group, also received same numbers of sessions (3 times) with the turned off device (Placebo). It should be mentioned that for the patients of both groups, in case of sense the pain, pethidine ampoule was used as a slow intravenous infusion and its amount was recorded for further evaluations. For each patient, the postoperative pain level will be assessed using the VAS ruler where a score of 0 means complete painlessness and 10 is considered the most severe pain and the amount of medication received will be evaluated based on the medical recordings during the hospitalization period. Laser irradiation was performed using a low-power laser device with a Wavelength of 905 nm, frequency of 150 Hz and an output power of 40 mW at a distance of 2 cm from the edges of the wound without opening the dressing with a radiation dose of 2 J/cm2 per minute. Laser irradiation time was calculated 8 minutes for each area. Also, during hospitalization and using the low-power laser device, local wound complications such as hematoma, abscess, erythema, and seroma in both groups will be examined and compared on a daily basis.

### Statistical Analysis

SPSS version 26 software was used for data analysis. First, descriptive statistics including mean, standard deviation, and median were used, and then independent t-test was used to compare the average pain intensity in two groups at different times. In order to compare the averages in one group, the paired t-test was used at different time intervals. To compare the qualitative data, Pearson’s chi-square test was used, and a P-value of less than 0.05 was considered a significant result.

### Ethical considerations

It should be noted that before starting the study, the informed consent and the study protocol was approved by the regional ethics committee with the ethical code IR.BMSU.BAQ.REC.1401.029 and Iranian Clinical Trials committee with IRCT code 20221018056225N1. Before starting the research, according to the Helsinki regulations, the objectives of the study were announced to the patients and included in a separate section of the informed consent form, and all patients entered the study by completing and writing the informed consent form. Also, in case of unwillingness at any stage of the study, the patient was free to withdraw from the study.

## Result

In the intervention group, 27 patients were men (54%) and 23 were women (46%), and in the control group, 25 patients were men (50%) and 25 were women (50%). There was no statistically significant difference between the sex distribution of the two control and intervention groups, so that the P-Value of Pearson Chi-Square test was obtained as 0.689. In the intervention group, the average age obtained was 52.3 years with a standard deviation of 8.24 and in the control group, the calculated average age was 50.2 years with a standard deviation of 8.81.The average age obtained in the intervention and control groups did not have a statistically significant difference with each other, and the P-value calculated through independent t-test was 0.204. In the intervention group, the average WBC obtained before the operation was 6.48*10^3^ Cells/μl/cu mm with a standard deviation of 1.8*10^3^ Cells/μl/cu mm and the average ESR and CRP obtained before the operation, it was calculated as 8.3mm/hr with a standard deviation of 3.6mm/hr and 5.6 with a standard deviation of 3.1 respectively. Also, the average WBC obtained 24 hours after the operation was 9.18*10^3^ Cells/μl/cu mm with a standard deviation of 2.69*10^3^ Cells/μl/cu mm and the average ESR and CRP obtained 24 hours after the operation, it was calculated as 34.92mm/hr with a standard deviation of 30.37mm/hr and 110.38 with a standard deviation of 38.17 respectively. On the third day after the operation, the average WBC was obtained 6.49*10^3^ Cells/μl/cu mm with a standard deviation of 2.69*10^3^ Cells/μl/cu mm and the average ESR and CRP were obtained 32.48 mm/hr with standard deviation 26.24 mm/hr and 32.3 with standard deviation 20.24 respectively. In the control group, the average WBC obtained before the operation was 6.64*10^3^ Cells/μl/cu mm with a standard deviation of 1.84*10^3^ Cells/μl/cu mm and the average ESR and CRP obtained before the operation, it was calculated as 7.74mm/hr with a standard deviation of 3.34mm/hr and 6.5 with a standard deviation of 3.2 respectively. the average WBC obtained 24 hours after the operation was 9.28*10^3^ Cells/μl/cu mm with the standard deviation of 2.83*10^3^ Cells/μl/cu mm and the average ESR and CRP obtained 24 hours after the operation were obtained 48.4 mm/hr with a standard deviation of 31.88 mm/hr and 115.56 with a standard deviation of 41.49 respectively. On the third day of the control group, the average WBC was obtained 6.78*10^3^ Cells/μl/cu mm with a standard deviation of 2.9*10^3^ Cells/μl/cu mm and the average ESR and CRP were obtained 34.3 mm/hr with a standard deviation of 25.9 mm/hr and 34.6 with a standard deviation of 18.4 respectively. The average of the investigated inflammatory factors in the intervention and control groups were not statistically significantly different from each other; So that comparing with the independent t-test, all the P-value numbers were greater than 0.05. In the intervention group, one case of abscess, four cases of seroma, and one case of hematoma, and in the control group, two cases of abscess, four cases of seroma, and one case of hematoma were observed, and there was no significant difference in the comparison; If comparing the above cases with the Pearson Chi-Square test, the P-Value number was more than 0.05. In the course of Comparison of pain intensity based on VAS in two groups(table1) In the first measurement of pain intensity in two groups, the average amount of pain in the intervention group was 7.22 cm with a standard deviation of 0.91 cm and in the control group it was 7.14 cm with a standard deviation of 1.01 cm. In the comparison between the two groups, there was no statistically significant difference and the P-Value based on the independent t-test was 0.678. But in the second measurement that took place 24 hours after surgery; The average pain intensity in the intervention group was 5.3 cm with a standard deviation of 1.03 cm and in the control group it was 6.3 cm with a standard deviation of 0.93 cm. At the this point of the time, the situation was different. Based on the independent t-test and in terms of pain intensity comparison, there was a significant difference between the average pain intensity in the two groups; So that this amount in the intervention group was significantly lower than the control group and the P-Value was calculated to be less than 0.001. At the third point of time i.e. 48 hours after surgery, the average pain intensity in the intervention group was 4.2 cm with a standard deviation of 1.03 cm and in the control group it was 5.3 cm with a standard deviation of 1.12 cm. In the comparison between the two groups, there was still a statistically significant difference in terms of pain intensity; So that in the intervention group it was significantly less than the control group and the P-Value was calculated less than 0.001. At the fourth time point, i.e. 72 hours after surgery, the average pain intensity in the intervention group was 3.66 cm with a standard deviation of 0.96 cm and in the control group was 4.3 cm with a standard deviation of 0.76 cm. Here too, in terms of comparison of pain intensity, there was a significant difference between the two groups, and the intensity of pain in the intervention group was significantly less than the control group; So that the P-Value was calculated to be less than 0.001. At the last time point, i.e. 96 hours after surgery, the average pain intensity in the intervention group was 2.44 cm with a standard deviation of 0.83 cm and in the control group it was 2.7 cm with a standard deviation of 0.81 cm. In the last comparison, as in the first time, there was no statistically significant difference between the intervention group and the control group in terms of pain intensity; So that the P-Value equal to 0.119 was calculated. In the course of comparison of pethidine consumption in two groups(table2); During the first 24 hours after surgery, the average consumption of pethidine in the intervention group was 53 mg with a standard deviation of 8.2 mg and in the control group was 54 mg with a standard deviation of 9.25 mg. There was no statistically significant difference between the control and intervention groups in terms of pethidine consumption in the first 24 hours after surgery, and the P-value was calculated as 0.569. During 24 to 48 hours after surgery, the average pethidine consumed in the intervention group was 29 mg with a standard deviation of 9.25 and in the control group 37.5 mg with a standard deviation of 12. 6 mg. In the comparison between the two groups, the P-value in this period was less than 0.001, which was significant. Between 48 and 72 hours after the surgery, the mean pethidine consumption in the intervention group was 10 mg with a standard deviation of 12.3 mg and in the control group 22.5 mg with a standard deviation of 16.9 mg and in this case Also, there was a statistically significant difference between the pethidine consumed in the two groups; So that this amount in the intervention group was lower than the control group and the P-Value was calculated lower than 0.001. But in 72 to 96 hours after surgery, the average consumption of pethidine in the intervention group was 5.5 mg with a standard deviation of 10.4 mg and in the control group 8.5 mg with a standard deviation of 11.9 mg was measured. There was no significant difference between the two groups, so the P-Value equal to 0.185 was obtained.

**Table 1.** Pain scores of patients in two groups.

|  | Case Group | Control Group | P-value |
| --- | --- | --- | --- |
| **VAS of 0 th. hour** | 7.22± 0.91 | 7.14 ± 1.01 | 0.678 |
| **VAS of 24 th. hour** | 5.30 ± 1.03 | 6.30 ± 0.93 | <0.001 |
| **VAS of 48 th. hour** | 4.20 ± 1.03 | 5.30 ± 1.12 | <0.001 |
| **VAS of 72 th. hour** | 3.66 ± 0.96 | 4.30 ± 0.76 | <0.001 |
| **VAS of 96 th. hour** | 2.44 ± 0.83 | 2.70 ± 0.81 | 0.119 |

**Table 2.** Opioid consuming of patients in two groups.

|  | Case Group | Control Group | P-value |
| --- | --- | --- | --- |
| **0-24 hour consume** | 53 ± 8.2 | 54 ± 9.2 | 0.569 |
| **24-48 hour consume** | 29 ± 9.2 | 37.5 ± 12.6 | <0.001 |
| **48-72 hour consume** | 10 ± 12.3 | 22.5 ± 16.9 | <0.001 |
| **72-96 hour consume** | 5.5 ± 10.4 | 8.5 ± 11.9 | 0.185 |

## Discussion

In this study, in terms of demographic data, the average age of the patients in the present study is 51.2 years, and in terms of gender distribution, 52% of the patients are men and 48% are women, which is compared to the study of Movasaghi et al. (1) average age 41.2 and the gender distribution of 57% of males against 43% of females and the study of Carvalho et al. (5) average age 47.1 and the study of Ojea et al. (10) average age 44.1; The patients of the present study are slightly older, but there was no significant difference between the groups in terms of average age and gender distribution in the present study, while most studies have ignored this issue. About inflammatory factors(WBC,ESR,CRP) in the current study; there was no significant difference between the two groups before and after surgery. This issue can also be interpreted in the short term and also only in the case of surgical site pain, there is no relationship with inflammatory factors, and in fact, inflammatory factors are more related to various post-operative intra-abdominal complications. In reviewing other studies; similar to the present study, the study by Movasaghi et al. shows that there is no significant relationship between inflammatory factors and laser therapy, but in the study by Ojea et al. (10) suggests the effects of low level laser therapy in reducing ESR. As mentioned, there was no significant difference between the intervention and control groups regarding the complications of the surgical site. Also, in the review of similar studies, no data was found in this regard. In this study, the VAS scale was used to calculate pain intensity, and based on the findings, the intensity of pain in the intervention group was slightly higher than the control group at the beginning of the study, but this difference was not statistically significant (P-Value=0.678); From this finding, it can be concluded that at the beginning of the study, the intensity of pain in both groups was almost the same, and the patients had a similar level of pain before any intervention, which can be the result of the results obtained at later time points. make it more reliable. After the start of the study, the intensity of pain decreased significantly in both groups and statistically, this decrease was significant in both groups; But the noteworthy point in this regard was the statistically significant difference in the comparison between the two groups; So that the average intensity of pain at 24-72 hours after the operation in the intervention group was much lower than the control group. (P-Value < 0.001). In terms of the patients’ need for opioid drugs, an investigation was also carried out and the average pethidine consumption in both groups was calculated and compared separately. Similar to the reduction of pain, the amount of pethidine consumption was significantly reduced in both groups separately in all time periods.Despite the fact that in the early hours of the study, the amount of pethidine consumed in the control group was higher than the intervention group; However, this was not statistically significant and the P-Value obtained in the first 24 hours was 0.569 respectively.However, in 72-24 hours after surgery, the difference between the mean pethidine consumed between the two groups was significant; So that in the study group, pethidine consumption was evaluated less than the control group. (P-Value<0.001)As the charts have shown; During the 96 hours of evaluation for both groups, the intensity of pain and the need for opioid drugs decreased significantly, but in comparison between the two groups, the average intensity of pain and the amount of opioid drug consumption in 72-24 hours after the operation in The intervention group was significantly lower than the control group.Considering that in the review of other studies, a completely similar case was not obtained, but in general, other studies, like the current study, have shown the effect of low-power laser therapy in reducing postoperative pain and reducing the need for painkillers. In the current study, a low-power laser with a wavelength of 905 nm and an energy density of 2 J/cm2 was used for 8 minutes and daily for up to 3 days. Compared to other studies; It has been shown that low-power laser therapy in visible to infrared wavelengths (650 to 905 nm) with an energy density of 0.2 to 54 J/cm2 for periods of 1 to 10 minutes and in single dose sequences, Every 6 to 24 hours up to twice a week and with periods of 1 day to 7 weeks can have significant effects in reducing pain after discectomy, inguinal hernia repair (5), oral carcinoma mucoepidermoid tumor surgery and Radiotherapy after that (6), breast plastic surgery (7), endodontic surgery (8), orthodontic treatment with fixed instruments and also reduced pain after bariatric surgery (10), acute and chronic neck pain (11), joint pain (12), after tibia fracture surgery (13), pain after elective cesarean surgery (14) and reduction of inflammatory factors, edema and tissue bleeding after injury and can have the same effects as drugs have a pain reliever like NSAIDs (15). Therefore, in this study, we have obtained significant results with low density and dosage protocol of laser therapy.

### Limitations and Suggestions

Researchers are always faced with limitations in their research, some of which show themselves even at the beginning of their work. On the other hand, unwanted variables that may be the result of special designs and methods used in research; often, they endanger the internal and external validity of the research in various ways. One should be aware that it is impossible to control or completely eliminate these types of factors in research. However, researchers try to reduce these factors as much as possible. predict and use all the necessary precautions in order to reduce them. In the current research, there were limitations in the method of conducting the study and collecting data, some of which are mentioned:

1. In this research, a questionnaire and VAS scale were used; As a result, some people may have refused to give real answers and gave unrealistic answers.
2. This research was conducted on a specific spectrum and a small number of patients in only one treatment center, and this issue makes it difficult to draw a general conclusion from the study and generalize its results.

It is suggested that similar studies with a higher sample size, in longer periods of time, on more surgeries and in other medical centers, and considering that in the present study, the follow-up of patients only in the initial 96 hours of hospitalization. It is also suggested that in order to implement these types of studies as best as possible, intervention research plans should be considered as a mandatory procedure in medical centers and be assisted in their implementation as accurately as possible.

## Conclusion

In this study, it was shown that the intensity of pain after midline laparotomy for elective surgeries of the upper and lower gastrointestinal tract has decreased significantly in both groups, but the significant point in the comparison between the two groups is that there is a significant difference between There was an intervention group and a control group; So that in 72-24 hours after the operation, the average intensity of pain in the intervention group was lower than the control group. The need for pethidine was not significantly different in the first 24 hours in the intervention and control groups, but in the 72-24 hours, a significant difference was observed between the two groups; So Pethidine consumption in the intervention group was lower than the control group.

## Data Availability

Data are available from corresponder author (contact via corresponder email or phone number) for researchers who meet the criteria for access to confidential data.

## Acknowledgments

The paper was issued from my thesis for Postgraduate education in general surgery. Hereby, we acknowledge the vice chancellor of Research and Technology Affairs of baqiyatallah University of Medical Sciences, Supervisor professor dr.hamed gholizadeh and Sincere thanks of dr.Raei for his collaboration.

---

**Figure 1.**
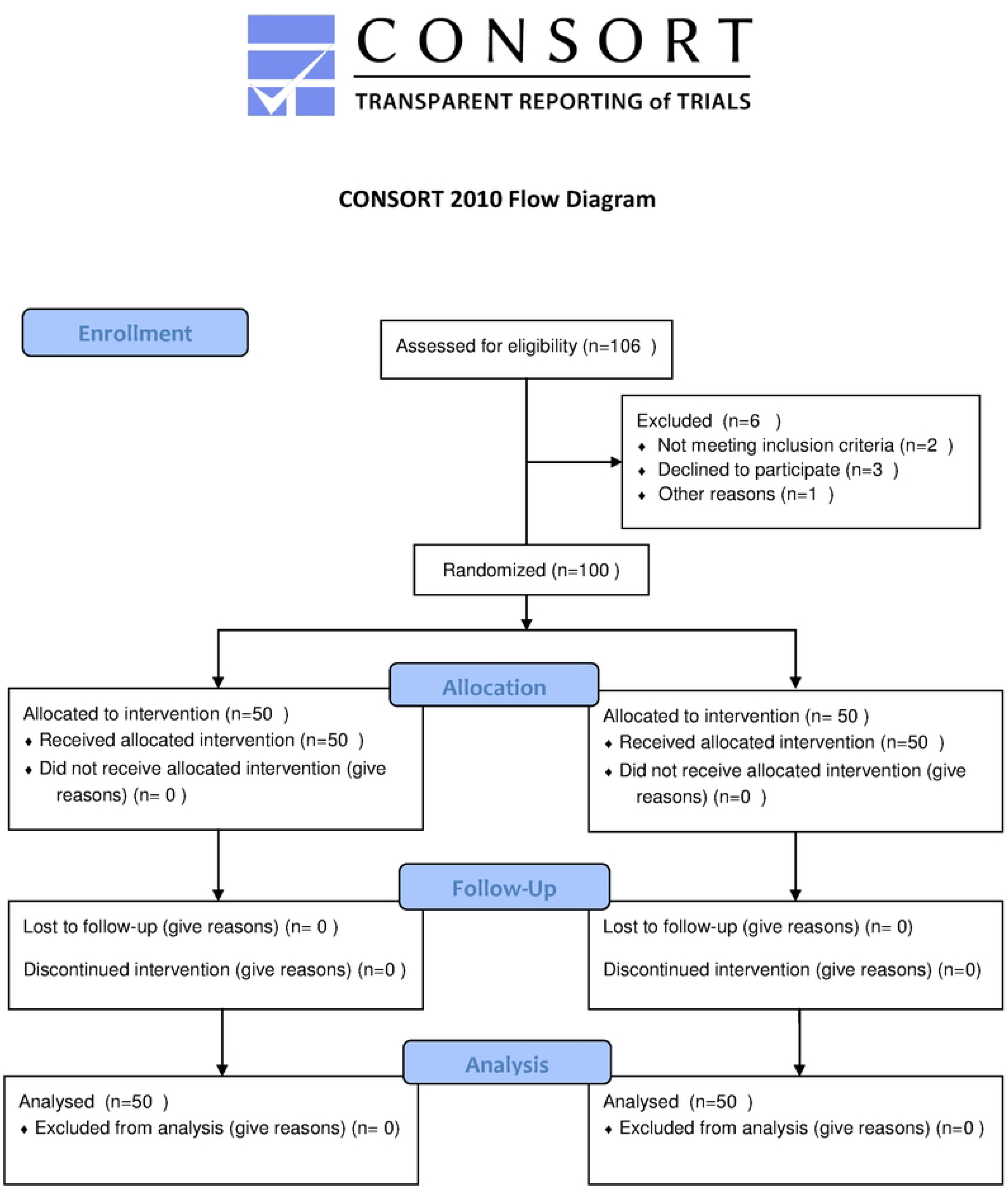

